# Wastewater-based detection of two influenza outbreaks

**DOI:** 10.1101/2022.02.15.22271027

**Authors:** Marlene K. Wolfe, Dorothea Duong, Kevin M. Bakker, Michelle Ammerman, Lindsey Mortenson, Bridgette Hughes, Peter Arts, Adam S. Lauring, William J Fitzsimmons, Emily Bendall, Calvin E. Hwang, Emily T. Martin, Bradley J. White, Alexandria B. Boehm, Krista R. Wigginton

## Abstract

Traditional influenza surveillance informs control strategies but can lag behind outbreak onset and undercount cases. Wastewater surveillance is effective for monitoring near real-time dynamics of outbreaks but has not been attempted for influenza. We quantified Influenza A virus RNA in wastewater during two active outbreaks on university campuses in different parts of the United States and during different times of year using case data from an outbreak investigation and high-quality surveillance data from student athletes. In both cases, the IAV RNA concentrations were strongly associated with reported IAV incidence rate (Kendall’s tau = 0.58 and 0.67 for University of Michigan and Stanford University, respectively). Furthermore, the RNA concentrations reflected outbreak patterns and magnitudes. For the University of Michigan outbreak, evidence from sequencing IAV RNA from wastewater indicated the same circulating strain identified in cases during the outbreak. The results demonstrate that wastewater surveillance can effectively detect influenza outbreaks and will therefore be a valuable supplement to traditional forms of influenza surveillance.

**Synopsis:** This study provides evidence that detection of Influenza A RNA in wastewater settled solids can be effectively used to track dynamics of influenza A outbreaks in a community.

## Introduction

Influenza surveillance is critical for determining the timing, location, and magnitude of outbreaks. Public health agencies use several combined data sources for influenza surveillance, including outpatient visits, hospitalizations, and clinical laboratory results. These methods can take 1-2 weeks or longer to detect increases in influenza activity^1^. Moreover, these surveillance systems generally characterize the beginning and peak of influenza season rather than their magnitude^2^, and only capture a small fraction of influenza illnesses^3^. People with milder illnesses, illnesses outside “typical” influenza season, or with limited access to medical care are generally unaccounted for. The COVID-19 pandemic has highlighted the need for more robust estimates and models of respiratory virus circulation, especially methods that can differentiate causative agents and identify outbreaks occurring outside typical seasonal patterns.

Wastewater-based epidemiology (WBE) is a surveillance technique that has recently been adopted widely. Other alternative surveillance approaches have been proposed to augment conventional influenza surveillance, including use of online activity^4^ and symptom data^3^; however, these tend to be non-specific to influenza and biased^3,5^. WBE targets genetic material from pathogens and is therefore highly specific; a number of teams have demonstrated that SARS-CoV-2 RNA levels in wastewater correlate with community incidence rates^6^ and identify the presence and trends of variants of concern^7,8^. Because fecal shedding of viruses that primarily infect the respiratory tract has been poorly characterized, WBE has not historically been used for these viruses. However, many respiratory viruses, such as RSV and influenza A, are shed and detectable in stool, and stool is not the only human contribution to wastewater ^9–12^. We have built on our successful deployment of WBE for COVID-19 surveillance to expand the use of these tools for RSV^13^, and in this study for the influenza A virus (IAV).

Measures to mitigate the transmission of COVID-19 dramatically limited the transmission of influenza over the first two years of the pandemic^14^, hindering efforts to investigate whether wastewater surveillance can be used for IAV. In fall of 2021, the University of Michigan (UM) in Ann Arbor, MI (USA) experienced a rapid increase in Influenza A (H3N2) cases, representing some of the first detected influenza activity in the United States since the start of the COVID-19 pandemic^15^. Given the outbreak’s novelty, several agencies–including members of our team– collaborated to collect high-resolution clinical data beyond usual surveillance^15^. In the spring of 2022, Stanford University in CA (USA) saw an increase in influenza A activity, as detected by comprehensive surveillance by the athletic department. These two outbreaks in different university communities, along with complimentary wastewater solids samples collected throughout the outbreaks, provided an opportunity to assess whether IAV RNA is detectable in wastewater solids and determine the relationship between wastewater concentrations and clinical data.

## Materials and Methods

### Assay choice for Influenza A

Droplet digital PCR was used to quantify a gene specific to Influenza A M1 gene using an assay by the United States Centers for Disease Control and Prevention (CDC)^16^. Multiple primer sets are available for this target from the CDC. Based on alignment with Influenza A sequences over the past 4 years in the United States primers INF A forward 1 and reverse 1 were chosen (see SI).

### Wastewater samples

Samples were collected from the City of Ann Arbor Wastewater Treatment Plant (AA WWTP). AA WWTP is a publicly owned treatment works that serves Ann Arbor, Michigan, USA, including the University of Michigan Ann Arbor campus. AA WWTP receives an inflow of 17 million gallons per day (MGD) from a sewershed with approximately 130,000 people, including students on campus and surrounding who utilize University Health Services. Further details on AA WWTP are provided in Kim et al.^17^ and in the SI. Samples were collected as part of a routine wastewater monitoring program, and samples between 4 Oct 2021 - 4 Dec 2021 were chosen for IAV analysis to match the time period of a known influenza A outbreak. During the study period, AA WWTP staff collected daily settled solids and 24 hour composite influent samples in sterile 50 ml conical tubes. These samples were stored immediately at 4°C and sent by courier to a laboratory at the University of Michigan where they were maintained at 4°C unless otherwise described; more details on sample storage are available in the SI.

Samples from Stanford University were collected from a manhole that accesses a large sewer main that conveys wastewater, from nearly 200 buildings on campus including student and faculty housing for 10,000 people. During the study period, 24-hour composite samples were collected and analyzed for IAV RNA 6 days per week as part of a routine monitoring program. A temperature controlled autosampler was set at 4°C and collected a raw wastewater sample every 30 minutes. The sample was then placed into an Imhoff cone to settle the solids; the solids were collected and stored at 4°C before processing on the same day. Samples between 1 Jan 2022 and 28 April 2022 during the period prior to and during an outbreak are included here.

### Sample preparation

Solids samples from both sites were dewatered by centrifugation followed by decanting and then resuspended in a buffer prior to direct extraction as described in Wolfe et al.^5^ Whole samples from the AA WWTP were pasteurized (60°C for 1 hour) prior to analysis to adhere to UM biosafety protocols. Influent samples from AA WWTP were concentrated using PEG to precipitate viruses as described by Flood et al a^18^. Additional details are available in the SI.

### RNA extraction

RNA extraction was performed on 10 replicate aliquots of dewatered settled solids or PEG pellet generated from the influent as described in Wolfe et al^6^. To remove inhibitors, samples were passed through a Zymo OneStep-96 PCR Inhibitor Removal Kit (Zymo Research, CA). Negative controls (water) and positive controls (BCoV spiked in DNA/RNA shield) were included for each extraction plate and 4 μL of Poly-A carrier RNA was added to the extraction positive controls prior to extraction.

### Droplet digital RT-PCR

Extracted RNA samples from both sites were immediately analyzed using two multiplex droplet digital RT-PCR (ddPCR) assays. One multiplex assay targeted the IAV M1 gene and SARS-CoV-2 nucleocapsid (N) and spike (S) genes; the IAV M1 gene alone is presented here. A second multiplex assay targeted pepper mild mottle virus (PMMoV) and bovine coronavirus (BCoV), and is described elsewhere^6^. PMMoV is a plant virus that is shed in human stool and abundant in wastewater globally^19,20^. It is used here to control for fecal strength and recovery of viral RNA^21^. BCoV was used as an exogenous recovery control; samples were required to have greater than 10% BCoV recovery. PCR negative and positive and extraction controls were included to ensure no contamination as described in Wolfe et al^6^. Additional details of assays, controls, and thermocycling conditions are available in the SI.

Each of the 10 replicate RNA extracted aliquots were run in their own well, and results from 10 wells were merged for analysis. Thresholding was done using QuantaSoft™ Analysis Pro Software (Bio-Rad, version 1.0.596). Concentrations for each RNA target were converted using dimensional analysis to concentrations of target copies/g dry weight of wastewater solids unless otherwise stated^22^. Dry weight of dewatered solids was determined by drying. Total error includes both error associated with the Poisson distribution and variability among merged replicates, and is reported as standard deviations.

### Sequencing Methods

IAV RNA from a settled solid sample collected at the AA WWTP during the peak of the outbreak (11 Nov, 2021) was sequenced to determine if IAV genotype in wastewater matched that identified in clinical specimens. RNA was converted to cDNA and then amplified with a SuperScript IV One-step RT-PCR kit with Platinum™ Taq High Fidelity DNA Polymerase using primers targeting the IAV HA sequence. The product was resolved with 1% agarose gel, and the putative HA amplicon was excised and amplified with primers targeting the HA1 subunit sequence. The product was purified using magnetic beads (AmpureXP, Beckman Coulter) and sequenced using the Sanger method with the HA1 subunit primers. Details of the sequence generation, including PCR primers and thermal cycler parameters are provided in the SI.

### Clinical data

For the University of Michigan outbreak, the daily total number of positive samples (hereafter “daily cases”) were provided by University Health Service at the University of Michigan (UHS). Data was collected by UHS as part of an outbreak investigation, and positive cases are presented as a function of specimen collection date^15^. The majority of cases reported to the county were captured by the UHS outbreak data (97%); we therefore calculated IAV incidence rates using the number of daily cases from UHS and the total sewershed population of 130,000. A 5-day centered, smoothed average was used in analysis to limit the impact of testing bias. These activities were conducted consistent with applicable federal law and determined not to be regulated by the IRB (HUM00209156).

At Stanford University, case data were provided from surveillance among student athletes. During the study period, any student athlete (approximately 900 total) presenting with flu-like symptoms was screened for COVID-19, and if negative, specimens were screened at the Stanford Virology Laboratory for IAV. Positive cases are presented as a function of specimen collection date. Student athletes live throughout campus and do not have preference for housing; therefore it is assumed that the fraction of athletes living in the sewershed is approximately equivalent to the fraction of campus served by the sewershed and that incident IAV cases among students on campus are proportional to the number of incident cases in the sewershed. Thus, IAV incidence rate was calculated using the number of daily cases from student athlete surveillance and the total student athlete population of 900. This study was approved by the Stanford University IRB (57358) with a waiver of informed consent.

### Statistical Analysis

For the outbreak periods at both UM and Stanford, primary analysis was performed using daily IAV M1 gene concentration along with 5-day smoothed incidence rate. This analysis was repeated with data normalized by PMMoV, and is included in the SI. We tested the null hypothesis that daily wastewater concentrations were not associated with 5 day smoothed incidence rates using Kendall’s tau as data were not normally distributed at either UM or Stanford (Shapiro-Wilk normality test, both p<0.05). The rate of change of smoothed IAV incidence rate with IAV wastewater concentrations was estimated using linear regression. Measurements below the detection limit were replaced with 500 copies/g (the approximate detection limit), and days with 0 clinical cases reported were replaced with 0.1 for analysis. All statistical analyses were implemented in Rstudio (version 1.3.1073).

## Results and Discussion

We found that IAV RNA in wastewater was correlated with IAV incidence rate at both UM and Stanford. The first clinical case was reported at UM on 6 Oct 2021. No more cases were reported until October 20; the outbreak then rose to a peak of 84 daily cases on 10 Nov 2021 (Fig 1, Fig S1). Measurements of the IAV M1 gene in archived wastewater were below the detection limit (approximately 500 copies/g) from October 4 until the first positive measurement on 21 Oct 2021 and then rose and peaked on 17 Nov 2021 before the end of the study in early December (Fig 1). IAV M1 gene concentrations ranged from non-detect to 2.63×10^4^ copies/g (median 1.03×10^4^ copies/g). Overall, 857 cases were reported by UHS during the study, accounting for 97% of cases reported at the county level and thus approximating the cases in the Ann Arbor sewershed (Fig S2). Positive and negative controls were positive and negative, respectively. BCoV recoveries were above 10% and PMMoV concentrations were within the expected range for each plant, suggesting acceptable recovery of RNA. Wastewater data are available publicly through the Emory Dataverse for UM (https://doi.org/10.15139/S3/TMOEDS) and through the Stanford Digital Repository for Stanford (https://purl.stanford.edu/mb859sw3898). Case data is available in Fig S1.

**Fig 1.**
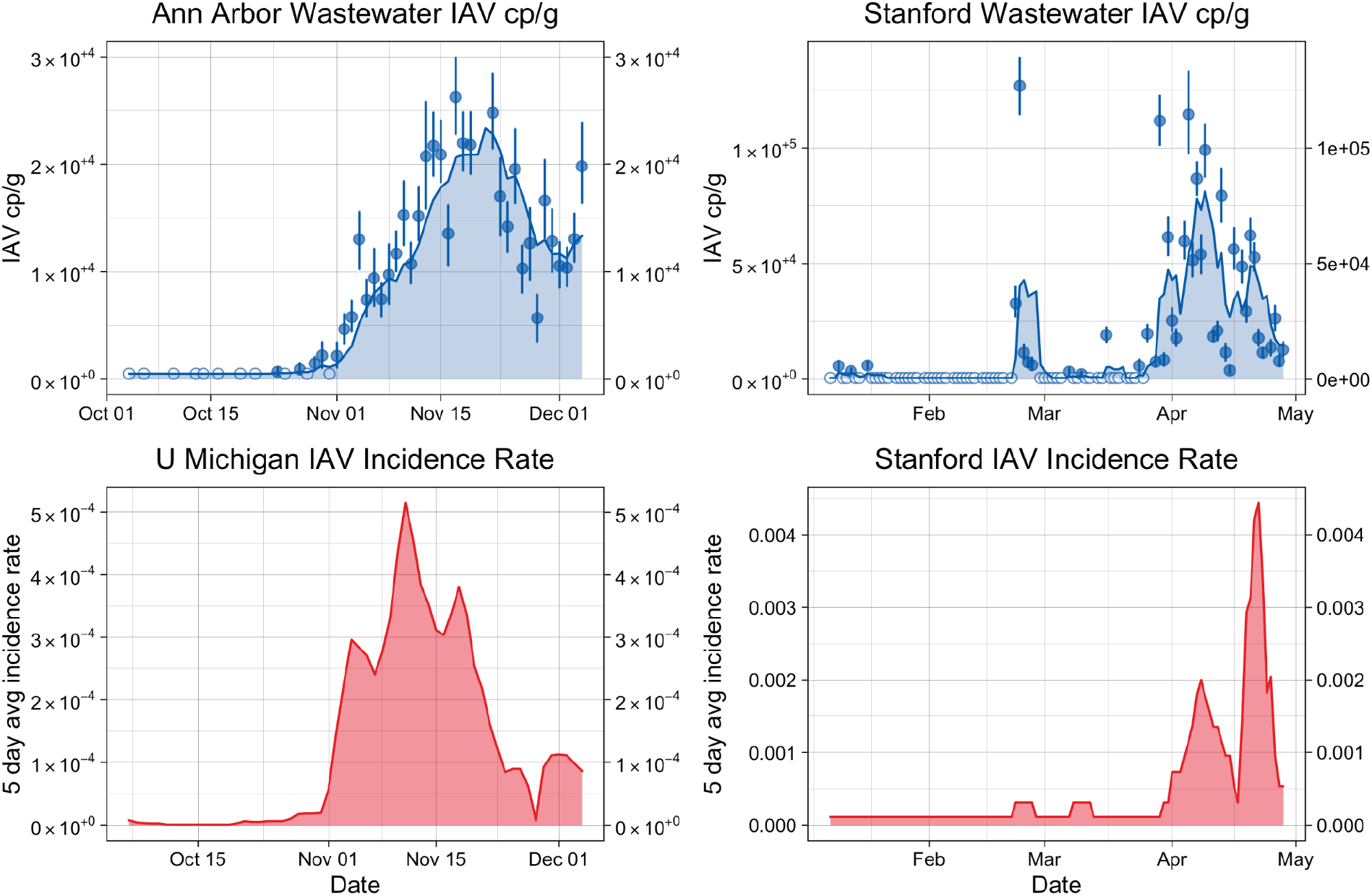
Time series of the University of Michigan and Stanford outbreaks, as shown in wastewater IAV concentrations and confirmed influenza A incidence rate. In the top row, points show daily IAV M1 RNA concentrations in copies/g dry weight of wastewater solids with error bars representing standard deviations of replicates as total errors from the ddPCR instrument software. Open circles indicate non-detect measurements. The area under the line represents the 5 day smoothed average copies/g IAV M1 RNA in wastewater. In the bottom row, the red area represents the 5 day smoothed number of reported cases and for UM the black line represents the 5 day smoothed positivity rate of clinical specimens.

The first clinical case of IAV infection in a Stanford athlete was reported on 22 Feb 2022, with only one other case reported until March 30, after which cases increased to a maximum of 7 per day on 19 Apr 2022 (Fig 1). Measurements of IAV M1 gene from a real time, daily monitoring program were below the detection limit when monitoring began with only a few positive measurements in January 2022 during a period without recorded cases after which there were increases in concentration seen starting on February 22 and again around 26 Mar 2022 (Fig 1). From the end of February, IAV RNA levels rose and then peaked at 1.15×10^5^ copies/g on 5 Apr 2022. Overall IAV M1 gene concentrations ranged from non-detect to 1.27×10^5^ copies/g (median non detect) and a total of 42 student athlete cases were reported. Positive and negative controls were positive and negative, respectively. BCoV recoveries were above 10% and PMMoV concentrations were within the expected range.

At both UM and Stanford, wastewater concentrations of IAV RNA closely reflect the IAV incidence rate (Fig 2). Daily IAV RNA concentrations were highly associated with both 5-day smoothed incidence rate (for UM tau = 0.58, p<10^−7^, N = 45, for Stanford tau = 0.67, p<10^−14^, N = 83). Results were not substantially different at either site when wastewater concentrations were normalized by an endogenous control (PMMoV) (see SI, Table S1, Fig S3). A 1 log_10_ increase in influenza A RNA in wastewater is associated with a 1.2 log_10_ increase in incidence rate at UM (r^2^ = 0.77, p <10^−14^, N = 45) and a 0.44 log_10_ increase in incidence rate at Stanford (r^2^ = 0.60, p <10^−16^, N = 94). A trimmed 992-base sequence (see SI) was generated from the 11 Nov AA WWTP sample and was identical to 212 IAV (H3N2) clinical samples that were sequenced during the 2021 IAV outbreak in Ann Arbor (see SI for sequence).

**Fig 2.**
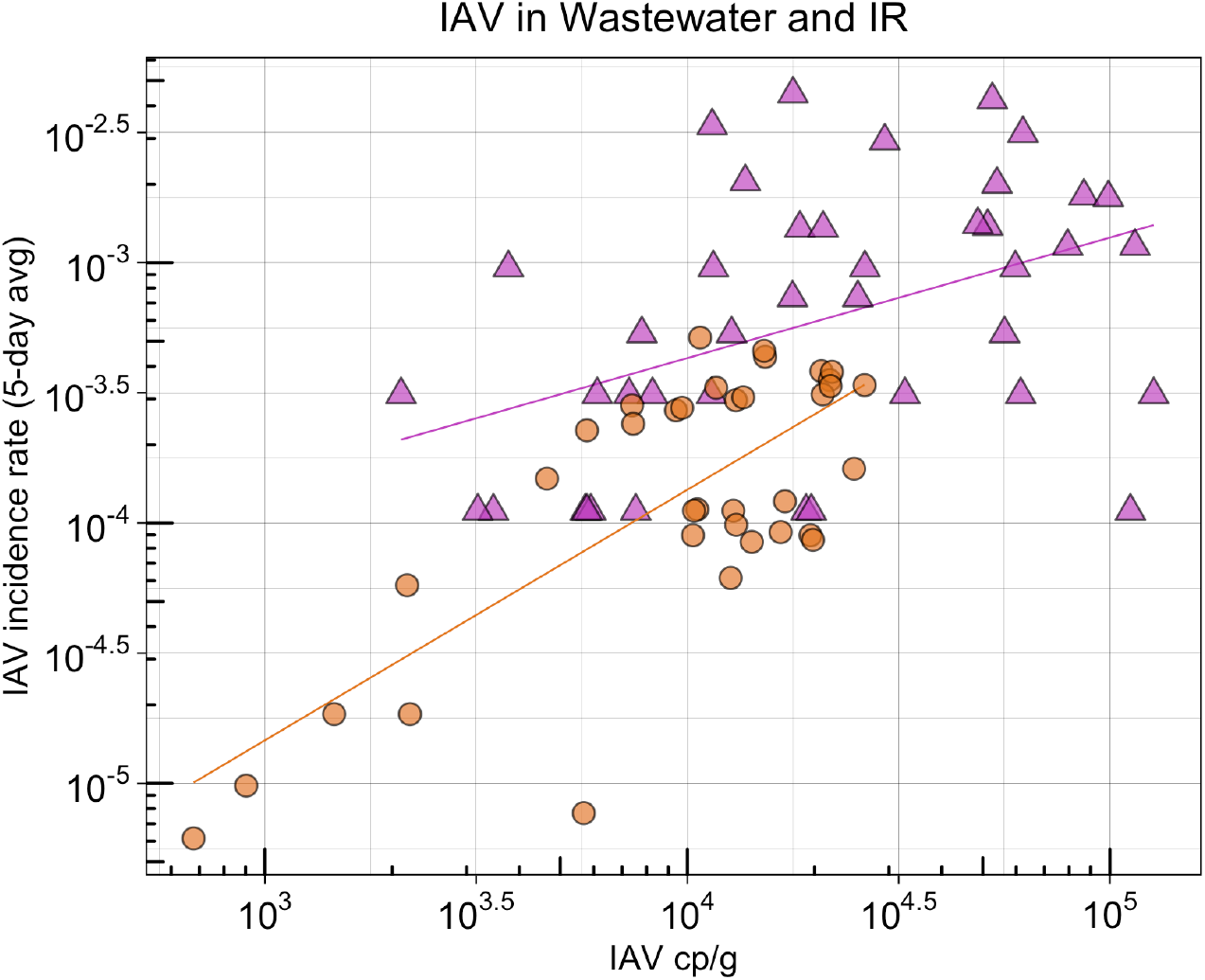
Association between positive daily Influenza A M1 gene concentration per gram dry weight and 5 day smoothed incidence rate for UM (orange circles) and Stanford (purple triangles). The line represents the linear association between the two measurements for each site.

The strength of the association between IAV RNA concentrations and clinical cases at both sites is similar to those reported for SARS-CoV-2 measured across eight publicly owned treatment works [Kendall’s tau value of 0.66 (p< 0.001) compared to 0.58 (p<0.001) and 0.67 (p<0.001) for IAV at UM and Stanford, respectively]. However, the linear relationship between wastewater measurements and incidence rate is different for SARS-CoV-2 and each IAV outbreak. A one log_10_ increase in the SARS-COV-2 RNA corresponded to a 0.59 log_10_ increase in incidence rate^6^ compared to a 1.2-log_10_ and 0.44-log_10_ increase in IAV incidence reported here for UM and Stanford, respectively. This observation could be due to differences in shedding of the virus between SARS-CoV-2 and IAV, differences in wastewater infrastructure and travel times, or differences in clinical testing and reporting. IAV RNA, like SARS-CoV-2 RNA, appeared at higher rates and at higher magnitudes in solids than in liquid wastewater in a comparison of four samples from UM (details in SI).

Previous research suggests that some viruses are highly concentrated in wastewater solids^23,24^. We compared concentrations of IAV RNA in four paired liquid influent and settled wastewater solids samples. We detected IAV RNA in all four of the solids samples and in only one of the four influent samples. IAV RNA concentrations were 10^3^-fold higher on a mass equivalent basis (Fig S4) in the solids sample that matched the positive influent sample. Our results from the paired wastewater influent and solids samples suggests that IAV RNA is more frequently detected in the solids fraction than the liquid fraction of wastewater. We therefore focused on settled solids in the study. These results suggest that IAV wastewater surveillance will be more sensitive and effective if sampling and measurements focus on the solid portion of municipal wastewater.

This work has several limitations - although the clinical data available from both UM and Stanford is higher quality than typical influenza surveillance data (due to an outbreak investigation at UM and active student athlete surveillance at Stanford), in both cases the wastewater catchment area does not exactly match the surveilled population. Thus, there is more work needed to clarify the relationship between IAV concentrations in wastewater and the number of cases in the contributing population. However, data from these two outbreaks together demonstrate that wastewater monitoring can effectively detect the rise and fall of cases during an outbreak of Influenza A. Longitudinal studies of shedding through infection with different IAV types and subtypes in a range of populations (e.g., vaccine status, age, etc.) would help contextualize these results for IAV and indicate if the same relationships between wastewater concentrations and incidence would be expected for the other influenza types and subtypes and in different communities. Additional research is also necessary to characterize the potential leading or lagging of wastewater compared to influenza symptom onset, testing, and reporting dates.

Wastewater surveillance of influenza could aid clinical surveillance, especially when traditional systems are not fully engaged (e.g. outside of typical flu season). Relative levels of different co-circulating influenza strains and types could be measured simultaneously to provide early identification of drifted strains not covered by the vaccine, a situation that would prompt additional public health guidance for testing and treatment^25^. Wastewater surveillance could also provide efficient monitoring for rare but impactful emergencies, such as introductions of new influenza strains at the animal-human interface with uncertain and potentially severe public health consequences^26,27^. Finally, distinguishing between influenza-like illnesses (ILIs) can be challenging; detection of these outbreaks were due largely to equipment that facilitated frequent testing with a 4-plex virus panel at UM, and active athlete surveillance at Stanford. This type of in-house clinical testing and surveillance is not widely used. Wastewater testing for a panel of IAV, SARS-CoV-2, and other viruses would give public health entities the ability to monitor several respiratory diseases in the community simultaneously and anticipate disease occurrence and strain on local healthcare systems. We anticipate wastewater surveillance will ultimately play an important role in improving the health system and public health preparedness for IAV and other respiratory viruses as it has done for SARS-CoV-2.

## Data Availability

All data produced in the present study are available upon reasonable request to the authors, UM wastewater data is available at https://doi.org/10.15139/S3/TMOEDS and through the Stanford Digital Repository for Stanford (https://purl.stanford.edu/mb859sw3898)

https://purl.stanford.edu/mb859sw3898

## Funding

CDC Foundation, Stanford University Provost’s Office, MDHHS, UMich COE Skunkworks We thank Miranda Delahoy (CDC), Kate Kusiak Galvin (UMich), Earl Kenzie (AAWWTP), Kyle Weinman (AAWWTP) for their contributions to and support of this work.

## Notes

### Competing Interest Statement

Dorothea Duong, Bridgette Hughes, and Bradley J. White are employees of Verily Life Sciences. Marlene K Wolfe previously provided consulting for Verily Life Sciences.

### Funding Statement

This study was funded by the CDC Foundation, Stanford University Provost's Office, MDHHS, UMich COE Skunkworks

### Author Declarations

The IRB of the University of Michigan waived ethical approval for this work (HUM00209156). This study was approved by the Stanford University IRB (57358) with a waiver of informed consent.

### Summary of Updates

This manuscript was updated to include a second outbreak of influenza and evidence from sequencing matching clinical and wastewater specimens.

