## Supplemental Information for "Wastewater-based detection of two influenza outbreaks"

### Supplementary Methods

**Sampling.** The University of Michigan Ann Arbor resumed in person classes for the 2021-2022 academic year and enrolled over 50,000 students in the Fall of 2021. Thus, the University of Michigan student body contributes significantly to the AA WWTP sewershed population. Stanford University samples were collected from an area that exclusively serves the campus population, which includes student athletes.

**Pre-analytical processing.** At the Ann Arbor Wastewater Treatment Plant (AA WWTP) thirty six settled solid samples from 10/28/2021 to 12/4/2021 were stored at 4°C at the time of sample collection, and prior to shipping for analysis on 12/8/21 were pasteurized and dewatered by centrifugation at 17,000 rpm for 2 minutes. An additional ten solids samples from 10/4/2021 to 10/27/2021 were prepared slightly differently; samples were centrifuged at 24,000xg (~14,000 rpm) for 30 minutes and the dewatered solids frozen at -80°C at the time of sample collection. Specimens were thawed and a 200-350 mg aliquot, removed and refrozen at -80°C prior to shipping for analysis. Four paired solids samples were provided not pasteurized to investigate any effect of pasteurization. Matching influent wastewater samples were selected for four dates to compare results in solids and liquids. Forty milliliters of each influent sample was concentrated to between 1.4 and 3.6 ml using PEG to precipitate viruses as described by Flood *et al* and concentrates were stored at -80°C until RNA extraction was performed[1]. Influent samples were not spiked with BCoV.

Only solids samples were processed from Stanford, and samples were processed on the same day a collection after storage at 4°C.

**Influenza A Assay**
For all samples, the following primers and probes as reported by the CDC were procured from Integrated DNA Technologies (IDT, San Diego, CA) and used for analysis[2]. For the probe, FAM, 6-fluorescein amidite was used as the fluorescent molecule with an internal ZEN quencher (proprietary from IDT) and a 3' Iowa Black FQ quencher.

| | |
|---|---|
| InfA For-1 | CAA GAC CAA TCY TGT CAC CTC TGA C |
| InfA Rev-1 | GCA TTY TGG ACA AAV CGT CTA CG |
| InfA Probe | 5'-/FAM/TGC AGT CCT /ZEN/ CGC TCA CTG GGC ACG/3IABkFQ/-3' |

**Droplet digital PCR**. For both assays, ddRT-PCR was performed on 20 µl samples from a 22 µl reaction volume, including 5.5 µl template RNA, 5.5 µl of One-Step RT-ddPCR Advanced Kit for Probes (Bio-Rad 1863021), 2.2 µl Reverse Transcriptase, 1.1 µl DTT and primers and probes at a final concentration of 900 nM and 250 nM respectively. Primer and probes for the Influenza A M1 gene, BCoV, and PMMoV were purchased from Integrated DNA Technologies (IDT, San Diego, CA). The AutoDG Automated Droplet Generator (Bio-Rad, Hercules, CA) was used to generate droplets and PCR was performed using Mastercycler Pro (Eppendforf, Enfield, CT). The following cycling conditions were used for the assay containing the Influenza A M1 gene: reverse transcription at 50°C for 60 minutes, enzyme activation at 95°C for 5 minutes, 40 cycles of denaturation at 95°C for 30 seconds and annealing and extension at 61°C for 30 seconds, enzyme deactivation at 98°C for 10 minutes then an indefinite hold at 4°C. The following conditions were used the the PMMoV/BCoV assay: reverse transcription at 50°C for 60 minutes, enzyme activation at 95°C for 5 minutes, 40 cycles of denaturation at 95°C for 30 seconds and annealing and extension at 55°C for 30 seconds, enzyme deactivation at 98°C for 10 minutes then an indefinite hold at 4°C. The ramp rate for temperature changes were set to 2°C/second and the final hold at 4°C was performed for a minimum of 30 minutes to stabilize droplets. Droplets were analyzed using the QX200 Droplet Reader (Bio-Rad). A well had to have over 10,000 droplets for inclusion in the analysis. All liquid transfers were performed using the Agilent Bravo (Agilent Technologies, Santa Clara, CA).

PCR positive controls for each target assayed on the plate were run in one well, and NTC  and negative extraction controls were run in 3 wells. Positive controls for Influenza A consisted of a gRNA mixed with Zeptometrix Influenza A controls. In order for a sample to be recorded as positive, it had to have at least 3 positive droplets. For the wastewater samples, three positive droplets corresponds to a concentration between ~500-1000 cp/g; the range in values is a result of the range in the equivalent mass of dry solids added to the wells. Results are reported as suggested in the EMMI guidelines[3].

**Clinical data.** UHS is the primary on-campus student healthcare facility. While it is open Monday-Saturday, hours on Saturday are limited to 3 hours. The raw case data reflects these patterns with fewer overall tests on Saturday and none on Monday. We also see an increase in total tests (positive and negative) on Mondays, likely due to the weekend buildup of symptoms.

The relationship between cases and wastewater data was also analyzed using data provided by Washtenaw County, as the county may capture cases that are outside UM and UHS care but within the boundaries of the AA WWTP. Since the majority of cases that were identified during the outbreak in the county were accounted for in the UHS data (97%), UHS data was used for analysis. Case data for UHS is aggregated based on both referral date (the date that a case record was opened), or onset date (the date the symptoms were first reported). Results of analysis using data provided by Washentaw County at the county level were similar to those from UHS data (Table S1).

Stanford University student-athletes were tested via the Sports Medicine Clinic on the Stanford campus. Influenza testing was available for symptomatic individuals seven days a week during the study period.

**IAV sequence recovered from Ann Arbor wastewater solids sample.**
Primary settled solids collected on November 11, 2021 were dewatered by centrifugation at 24000xg for 30 minutes at 4°C. A 58.2 mg sample of dewatered solid was homogenized in 1200 µL DNA/RNA shield (Zymo) with 0.5 mm silica/zirconia beads (Biospec) using the Mini-Beadbeater-96 (Biospec). Total nucleic acid was extracted from the homogenate using the Chemagic viral DNA/RNA 300 kit H96 (Perkin Elmer). The following primer set was used to target an IAV HA sequence:

Bm-HA-1TATTCGTCTCAGGGAGCAAAAGCAGGGG
Bm-NS-1TATTCGTCTCAGGGAGCAAAAGCAGGGTG

The following thermal cycler parameters were employed: 94°C for 2 min followed by forty cycles of 94°C for 30 s, 54°C for 30 s, and 68°C for 3min, followed by 68°C for 5 min and then held at 10°C.
The product was resolved on a 1% agarose gel, and the putative HA amplicon found at 1700 bp was excised from the gel and extracted using the GeneJet Gel Extraction Kit (Thermo Scientific). The extract was amplified using primers targeting the HA1 subunit sequence:

HA1_fwdATGAAGACTATCATTGCTTTGAGC
HA1_revTCTGGTTTGTTTCTCTGGTACA

The following thermal cycler parameters were employed: 50°C for 60 min and 94°C for two minutes followed by forty cycles of 94°C for 30 s, 50.4°C for 30 s, and 68°C for 3 min, followed by 68°C for 5 min.

The product was purified using a magnetic bead clean-up kit (AmpureXP, Beckman Coulter) and was sequenced using the Sanger method (Eurofins) with the HA1 subunit primers. Forward and reverse reads were trimmed by visualizing for good sequence chromatograms. Reads were concatenated using DNASTAR:Seqman Ultra version 17.3.0 (61) software and a fasta file was generated. Processed sequences were aligned using Augur to A/Cambodia/e0826360/2020, as well as clinical sequences from the 2020/2021 IAV outbreak in Ann Arbor, MI.

The final trimmed sequence was as follows:

GAGCAACATTCTATGTCTTGTTTTCGCTCAAAAAATACCTGGAAATGACAATAGCACGGCAACGCTGTGCCTTGGG
CACCATGCAGTACCAAACGGAACGATAGTGAAAACAATCACAAATGACCGAATTGAAGTTACTAATGCTACTGAG
TTGGTTCAGAATTCATCAATAGGTGAAATATGCGGCAGTCCTCATCAGATCCTTGATGGAGGGAACTGCACACTAA
TAGATGCTCTATTGGGGGACCCTCAGTGTGACGGCTTTCAAAATAAGGAATGGGACCTTTTTGTTGAAAGAAGCA
GAGCCAACAGCAACTGTTACCCTTATGATGTGCCGGGTTATGCCTCCCTTAGGTCACTAGTTGCCTCATCCGGCACA
CTGGAGTTTAAAAATGAAAGCTTCAATTGGACTGGAGTCAAACAAAACGGAACAAGTTCTGCGTGCATAAGGGGA
TCTAGTAGTAGTTTTTTTAGTAGATTAAATTGGTTGACCAGCATAAACAACATATATCCAGCACAGAACGTGACTAT
GCCAAACAAGGAACAATTTGACAAATTGTACATTTGGGGGGTTCACCACCCGGATACGGACAAGAACCAAATCTC
CCTGTTTGCTCAATCATCAGGAAGAATCACAGTATCTACCAAAAGAAGCCAACAAGCTGTAATCCCAAATATCGGA
TCTAGACCCAGAATAAGGGATATCCCTAGCAGAATAAGCATCTATTGGACAATAGTAAAACCGGGAGACATACTTT
TGATTAACAGCACAGGGAATCTAATTGCTCCTAGGGGTTACTTCAAAATACGAAATGGGAAAAGCTCAATAATGA
GATCAGATGCACCCATTGGCAGATGTAAGTCTGAATGCATCACTCCAAATGGAAGCATTCCCAATGACAAACCGTT
CCAAAATGTAAACAGGATCACATACGGGGCCTGTCCCAGATATGTTAAGCAAAGCACCCTGAAATTGGCAACAGG
AATGCGAA

**Supplementary Results**

When daily IAV RNA concentrations were normalized by PMMoV to correct for changes in RNA extraction efficiency, differences in fecal strength, or degradation from storage or treatment conditions [4], results were similar for both UM (tau = 0.62, $p<10^{-9}$) and Stanford (tau = 0.55, $p<10^{-11}$, Fig S3, Table S1).

**QA/QC.** All positive and negative controls were positive and negative respectively for both UM and Stanford sites. BCoV recoveries were above 10% for all samples (with the exception of influent samples, where BCoV was not used as a process control, as described in the methods). PMMoV concentrations in solids were in the expected range reported in previous work for both locations, and similar from sample-to-sample at each site, suggesting that RNA was recovered efficiently and consistently during extraction (Fig S3) [5]. The lowest detectable concentration in solids, assuming 3 droplets were observed, is approximately 500 copies/g, but varies slightly from sample-to-sample depending on the mass of solids added to DNA/RNA Shield.

**Pasteurization**. A subset of 4 paired pasteurized and unpasteurized samples from UM did not suggest a clear effect of pasteurization (Fig S5). Of the 4 samples, 3 paired samples were positive and in similar concentrations, and one paired sample was negative for both treatments. Therefore, we pasteurized samples given the benefit that the work with pasteurized solids could then be conducted in a biosafety level 2 facility.

**Positivity Rate:** For UM, positivity rate was also available and analysis was repeated for positivity rate. For the association between daily IAV cp/g and 5 day averaged positivity rate, tau = 0.62, $p<10^{-8}$. The relationship between IAV RNA and clinical positivity rate at UM indicates that a 1 $\log_{10}$ increase in influenza A RNA is associated with a 0.9 $\log_{10}$ increase in IAV positivity rate at UM ($r^2 = 0.76$, $p <10^{-14}$, N = 45). Positivity rate data was not available for Stanford.

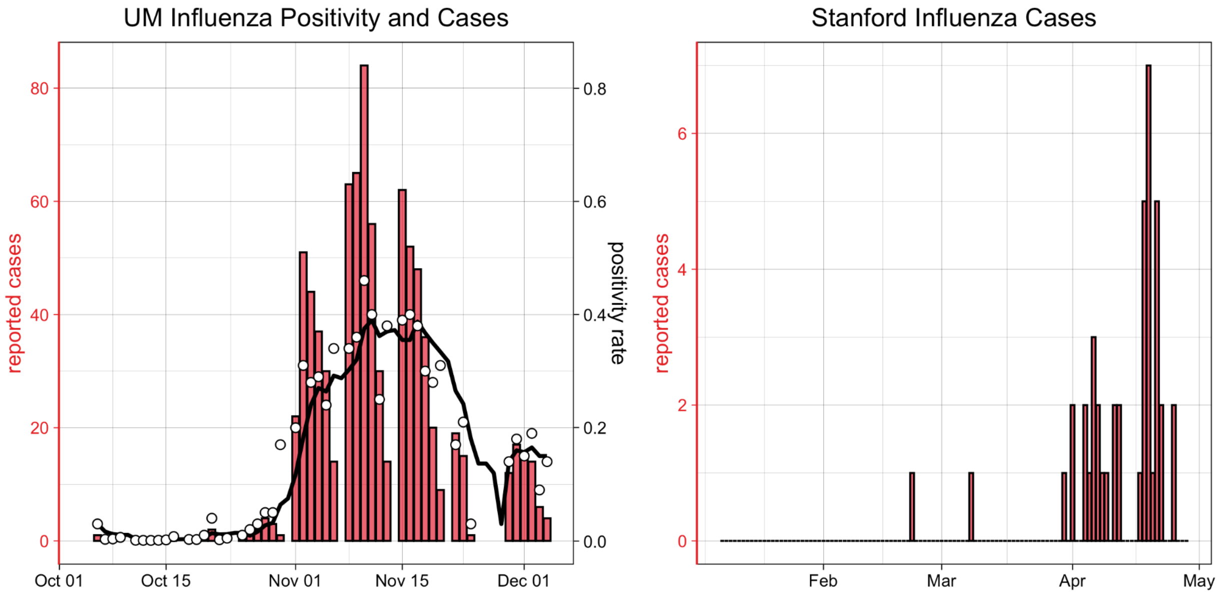


Fig S1. University of Michigan and Stanford University Influenza A clinical data. L) UM University Health Services clinical positivity rate data daily (points) and 5-day smoothed (line), alongside new cases (columns), and R) Stanford new daily cases (columns).

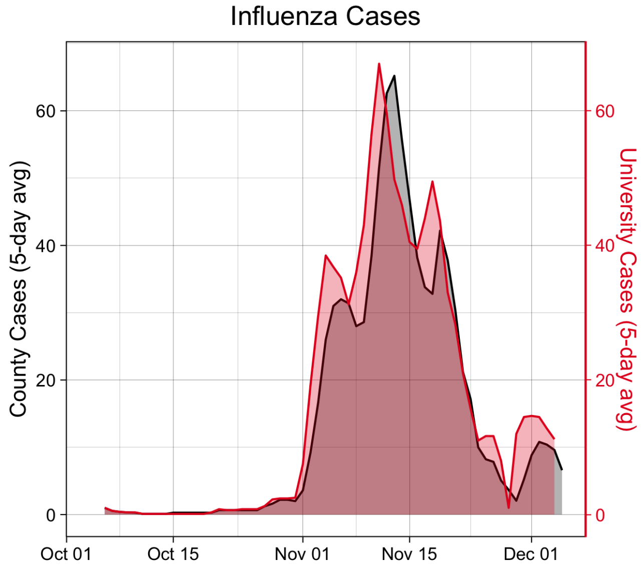


Fig S2. University of Michigan outbreak from 10/6/21 - 12/4/21 as shown in both UHS and Washtenaw County case data. The area under each curve represents the daily 5-day smoothed average of reported cases by test and referral date. A total of 887 cases were reported at county level by referral date and 857 cases were reported at UHS by test date. Thus, UHS data are estimated to capture 97% of reported cases in the outbreak period.

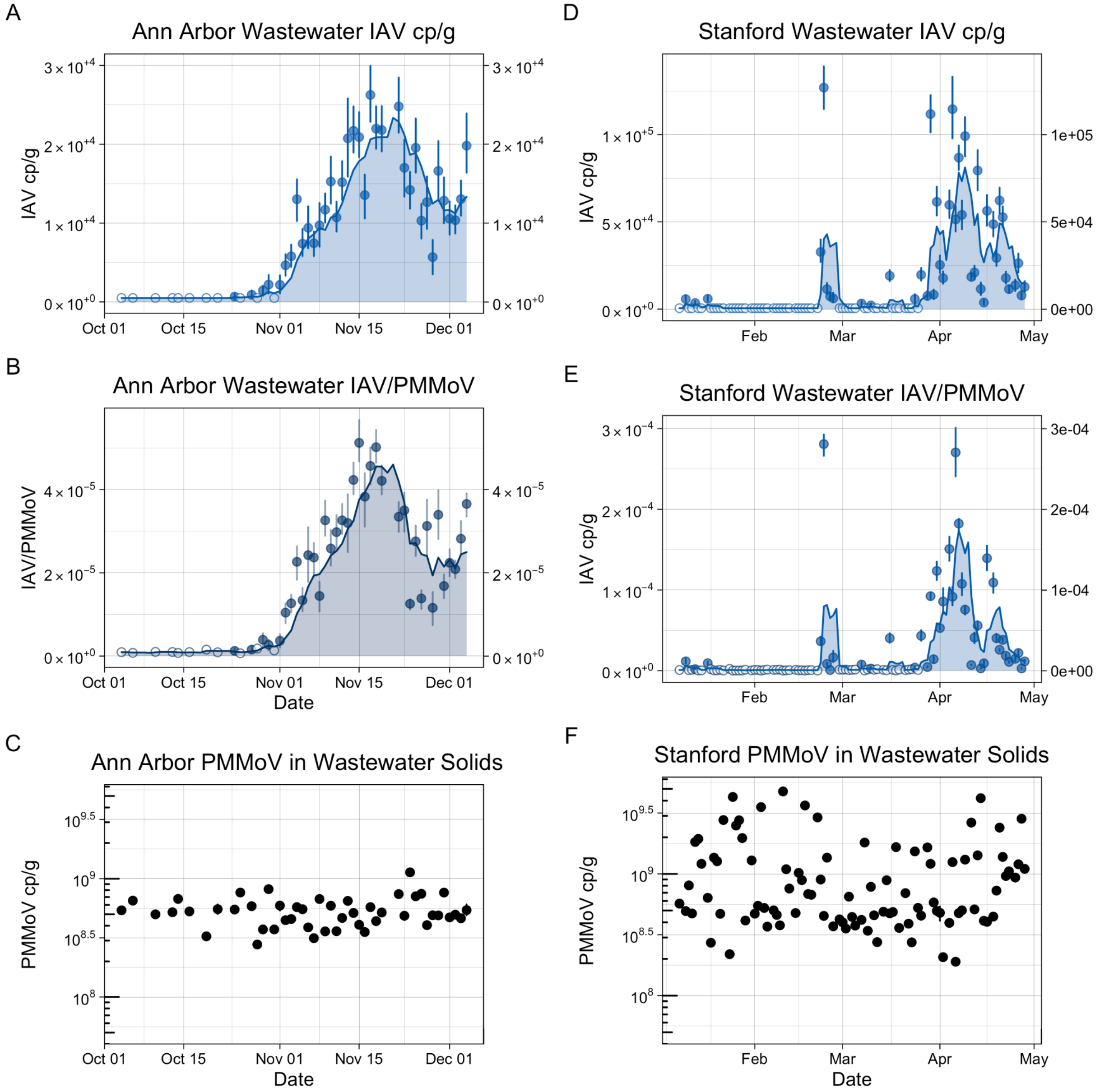


Fig S3. Time series of the University of Michigan outbreak, with IAV and PMMoV measured in wastewater. In panel A, points show daily IAV M1 RNA concentrations in copies/g dry weight of wastewater solids with error bars representing standard deviations of replicates as total errors from the ddPCR instrument software. Open circles indicate non-detect measurements. The area under the line represents the 5 day smoothed average copies/g IAV M1 RNA in wastewater. Panel B shows IAV normalized by PMMoV in the same manner. Panel C shows PMMoV RNA concentration in copies/g dry weight over time. Error bars are shown but are in most cases too small to visualize.

| Wastewater Variable | Case Variable | tau | p-value | df |
|---|---|---|---|---|
| AA WWTP IAV cp/g | UHS test positivity rate | 0.62 | 2.97E-09 | 44 |
| AA WWTP IAV cp/g | UHS incidence rate | 0.58 | 2.63E-08 | 44 |
| AA WWTP IAV cp/g | WC Onset incidence rate | 0.45 | 3.67E-05 | 44 |
| AA WWTP IAV cp/g | WC Ref incidence rate | 0.60 | 6.59E-09 | 44 |
| AA WWTP IAV/PMMoV | UHS positivity rate | 0.64 | 5.22E-10 | 44 |
| AA WWTP IAV/PMMoV | UHS incidence rate | 0.62 | 1.33E-09 | 44 |
| AA WWTP IAV/PMMoV | WC Onset incidence rate | 0.50 | 3.50E-06 | 44 |
| AA WWTP IAV/PMMoV | WC Ref incidence rate | 0.65 | 1.83E-10 | 44 |
| Stanford IAV cp/g | Stanford Athlete incidence rate | 0.67 | 4.32E-15 | 93 |
| Stanford IAV/PMMoV | Stanford Athlete incidence rate | 0.54 | 7.86E-12 | 93 |

Table S1. Kendall's tau for the relationship between wastewater measurements both in terms of copies/g dry weight and normalized by PMMoV, and clinical IAV cases as measured by University Health Services (UHS) and Washtenaw County (WC).

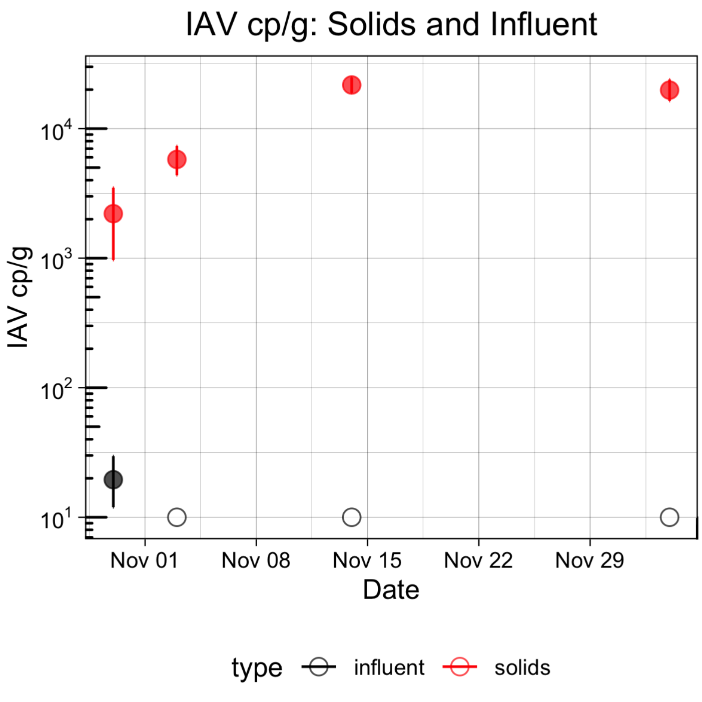

Fig S4. Comparison of paired solids and liquids samples. Open circles indicate that Influenza A was not detected. Points show RNA concentrations in copies/g dry weight of wastewater solids with error bars representing standard deviations of replicates as total errors from the ddPCR instrument software.

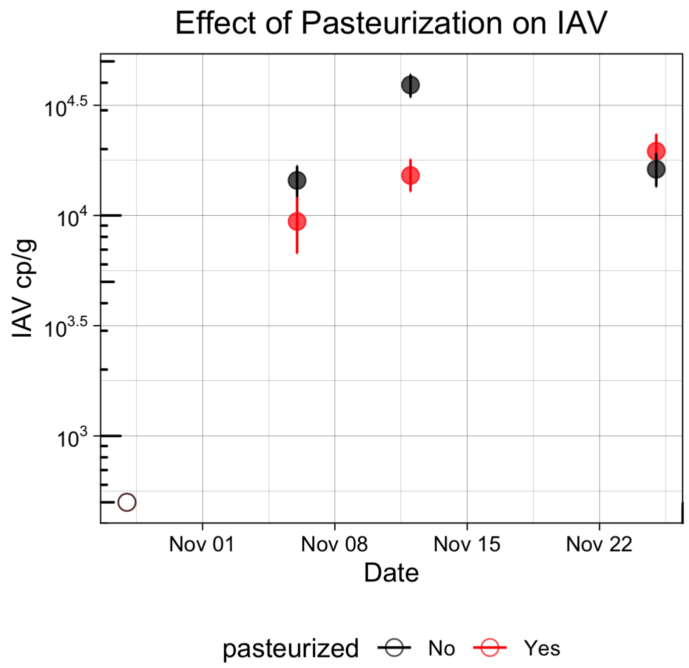


Fig S5. Comparison of paired pasteurized and unpasteurized solids samples. Open circles indicate that Influenza A was not detected. Points show RNA concentrations in copies/g dry weight of wastewater solids with error bars representing standard deviations of replicates as total errors from the ddPCR instrument software.

This paper has been previously submitted to medRxiv, a preprint server for Health Sciences. The preprint can be cited as:
